# Aspirin Use is Associated with Decreased Mortality in Patients with COVID-19: A Systematic Review and Meta-analysis

**DOI:** 10.1101/2021.07.08.21260236

**Authors:** Pramod Savarapu, Nischit Baral, Govinda Adhikari, Maxwell Akanbi, Basel Abdelazeem, Sakiru O. Isa, Ashiya Khan, Maham Ali, Sravanthi Jenumula, Kavitha Kesari, Arvind Kunadi

## Abstract

**Background:** Novel Corona Virus Disease 2019 (COVID-19) has resulted in more than three and half million deaths worldwide as of June 6, 2021. The role of aspirin in prevention of COVID-19 mortality has not been much studied. We aimed to study the relationship between aspirin use and covid-19 mortality.

**Methods:** We searched PubMed, MEDLINE, EMBASE, and Cochrane database for studies from January 2019 till June 6, 2021 with inclusion criteria of RCT, Cohort study, studies reporting mortality, and comparison studies on aspirin versus non-aspirin. Statistical analysis was done with Review Manager 5.4 statistical software using the inverse variance method. We assessed the pooled hazard ratio (HR), and 95% confidence interval using the random effect model and I-squared test was used to determine statistical heterogeneity.

**Results:** We included five retrospective cohort studies which met our inclusion criteria with total of 14065 participants in both groups. There were 6797 participants in the aspirin group and 7268 participants in the non-aspirin group. Our results show that the use of aspirin was associated with 53% decrease in mortality compared to non-aspirin in patients with COVID-19 (adjusted HR 0.47, 95% CI 0.35-0.63, P< 0.001, I^2^= 47%). In the analysis restricted to patients hospitalized for COVID-19, the use of aspirin was associated with a 49% reduction in the risk for in-hospital mortality (adjusted HR 0.51, 95% CI 0.33-0.80, P = 0.004, I^2^= 39%).

**Conclusions:** Our results show that aspirin is associated with decrease in both overall mortality and in-hospital mortality in patients with COVID-19.

## 1. Introduction

COVID-19 pandemic has resulted in more than three and half million deaths worldwide as of June 6, 2021. Although most patients with COVID-19 will have mild symptoms, the mortality rate for hospitalized patients is still high. As per a recent study, the overall mortality rate of COVID-19 patients (including both in-hospital and out-of-hospital) was 17.1% and mortality rate in hospitalized patients was 11.5%^1^. Immune dysregulation with systemic inflammation (especially IL-6) as well as thrombosis has been proposed a one pathway in the pathogenesis of severe COVID-19. Aspirin (acetylsalicylic acid) through its anti-inflammatory, antithrombotic with immunomodulatory effects may theoretically improve outcomes in patients with severe COVID-19^2^. We included more recently published studies in our meta-analysis to study the association between low-dose aspirin and mortality in patients with COVID -19.

## 2. Methods

### 2.1 Protocol and Registration

We did not register our study due to the need to submit the manuscript early for publication.

### 2.2 Search Strategy

We included Randomized control trials (RCTs), quasi experimental and cohort studies (including both prospective and retrospective cohort studies), that reported adjusted risk ratios of the effect of aspirin on mortality, including in-hospital mortality in patients with COVID-19. We searched both published and unpublished manuscripts and conference abstracts which fulfilled our eligibility criteria using appropriate mesh terms as described in the supplementary material. We searched PubMed, Google scholar and Clinical trials.gov from inception until May 6, 2021 to identify studies that investigated the effect of low dose aspirin on mortality among adults hospitalized for COVID-19, with no language restrictions. The search terms used are (aspirin OR acetylsalicylic acid) AND (COVID -19) AND (mortality), with a 5-year filter.

### 2.3 Eligibility Criteria

Eligible studies compared aspirin with no aspirin use in patients with COVID-19 and reported events of mortality using appropriate definition. We excluded studies that did not compare aspirin with no aspirin and didn’t report on mortality in COVID-19 patients. We did not include case reports, case series, and review articles in our meta-analysis.

### 2.4 Study Design

The Preferred Reporting Items for Systematic Reviews and Meta-Analyses (PRISMA) statement for reporting systematic reviews as recommended by the Cochrane Collaboration was followed in this systematic review^3^.

### 2.5 Data collection process

Search results were saved in EndNote version X9 (Developer: Clarivate analysis) files and transferred into Covidence software^4^. We extracted the data manually through full text review.

### 2.6 Selection Process

Two reviewers independently performed the title and abstract screening and full text screening. Conflicts were resolved through consensus and if not resolved, third author resolved the conflict.

### 2.7 Data Items

All the studies were compatible to each outcome domain. All the studies compared aspirin with no aspirin in COVID-19 patients. The outcome of mortality and/or in-hospital mortality was reported in all studies.

### 2.8 Methodical quality assessment

We used Newcastle-Ottawa Scale for assessment of quality of included studies. This scale assigns a maximum of nine points to each study. Score of four for selection and assessment of exposure, two for comparability, and three for assessment of the outcome. If a study receives a score of six or higher, it is considered a high-quality publication with low risk of bias.

### 2.9 Measure of Outcome

The outcome of interest was all-cause mortality (regardless of in or out of hospital) and in-hospital mortality.

### 2.10 Effect measure

We used hazard ratio for the effect measure.

### 2.11 Statistical analysis

Meta-analysis was done with Review Manager 5.4 statistical software using the inverse variance method. We assessed the pooled hazard ratio (HR), and 95% confidence interval using the random effect model. I^2^ statistic was used to assess heterogeneity. We did not perform meta-regression when heterogeneity was high and just reported the heterogeneity.

### 2.12 Sensitivity analysis

Sensitivity analysis was performed and showed no any change in the outcomes.

## 3. RESULTS

### 3.1 Study selection

We identified 120 articles from rom PubMed/MEDLINE, 683 articles from Embase and three articles from Web of Science. 521 articles were filtered out based on study design and criteria. Then 155 duplicate studies were removed. We identified 130 studies for full text assessment. Finally, 125 articles were removed for not meeting eligibility. Final qualitative and quantitative analysis was done with five studies (Figure 1).

**Figure 1:**
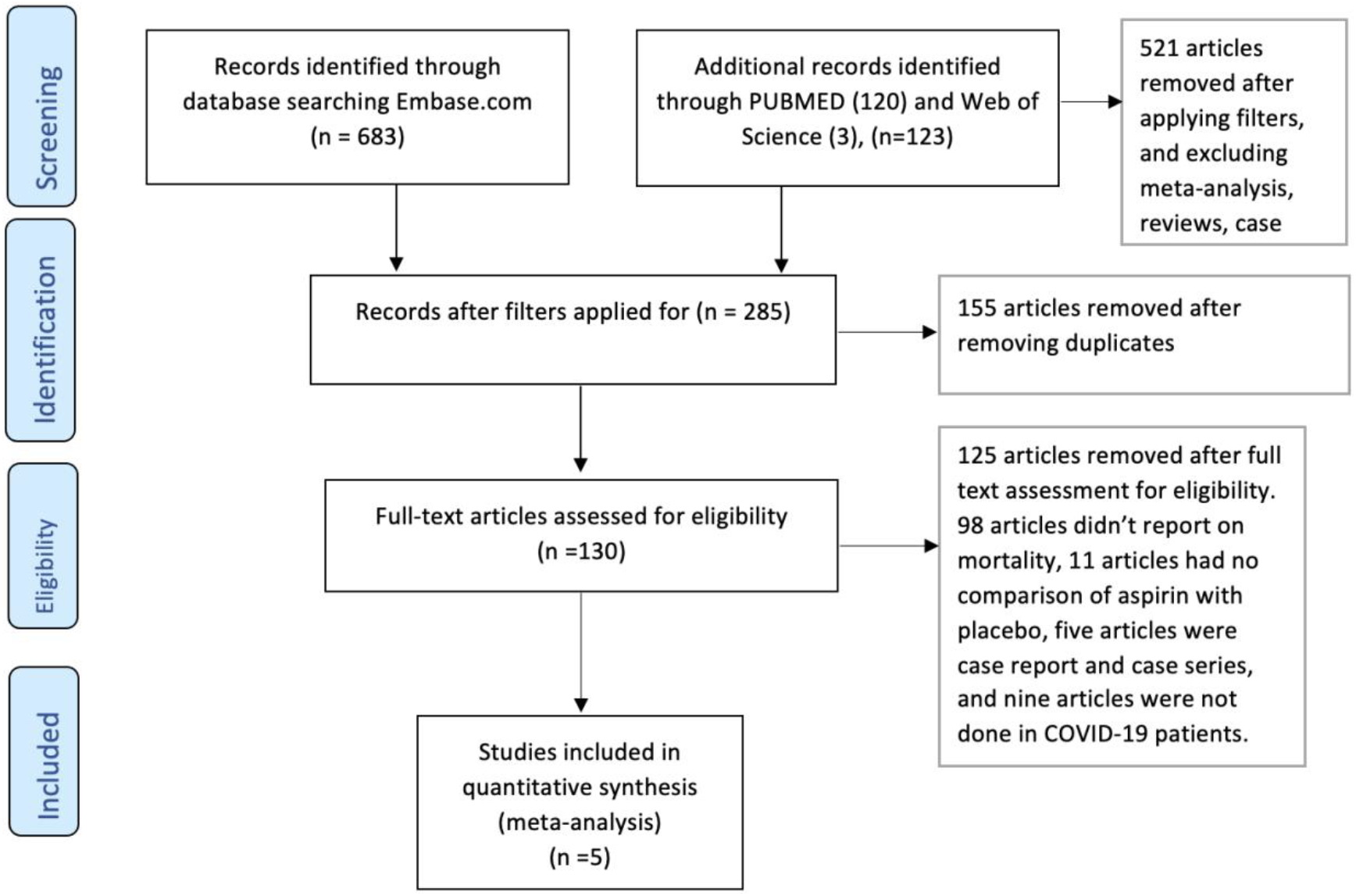

### 3.2 Baseline Characteristics of Studies

The baseline characteristics of included studies were extracted and has been listed in table 1. Of the five retrospective cohort studies included in our meta-analysis, all were observational studies, and four of the five studies were on hospitalized patients, and one included the general VA population in the US^3-7^. Apart from the Study by Yuan et al. which reported adjusted odds ratio, other studies reported adjusted HR^4^. To reduce confounding, included studies utilized multivariable-adjusted models, and (or) propensity matching. Table 1.

**Table 1.** Baseline characteristics of included studies.

| **ID** | **Country** | **Design** | **No of participants**<br>**(Aspirin vs non-Aspirin)** | **Study Population** | **Outcome** |
| --- | --- | --- | --- | --- | --- |
| Chow 2020 | USA | Retrospective | 98/314 | COVID Hospitalized | In hospital Mortality |
| Yuan 2020 | China | Retrospective | 52/131 | COVID Hospitalized | In hospital Mortality |
| Liu 2021 | China | Retrospective | 28/204 | COVID Hospitalized | 30- and 60-day Mortality |
| Meizlish 2021 | USA | Retrospective | 319/319(70/70) | COVID Hospitalized | In hospital Mortality |
| Osborne 2021 | USA | Retrospective | 6300/6300 | COVID Outpatient | 14/30-day Mortality |

### 3.3 Quality of included studies

The results of the Newcastle-Ottawa scale for assessment of quality of included studies showed that a four of the five studies had score of 6 and one had a score of 4^11^. This signifies that the quality of these 4 studies was high but one of the studies had low quality. (Table 2).

**Table 2.**
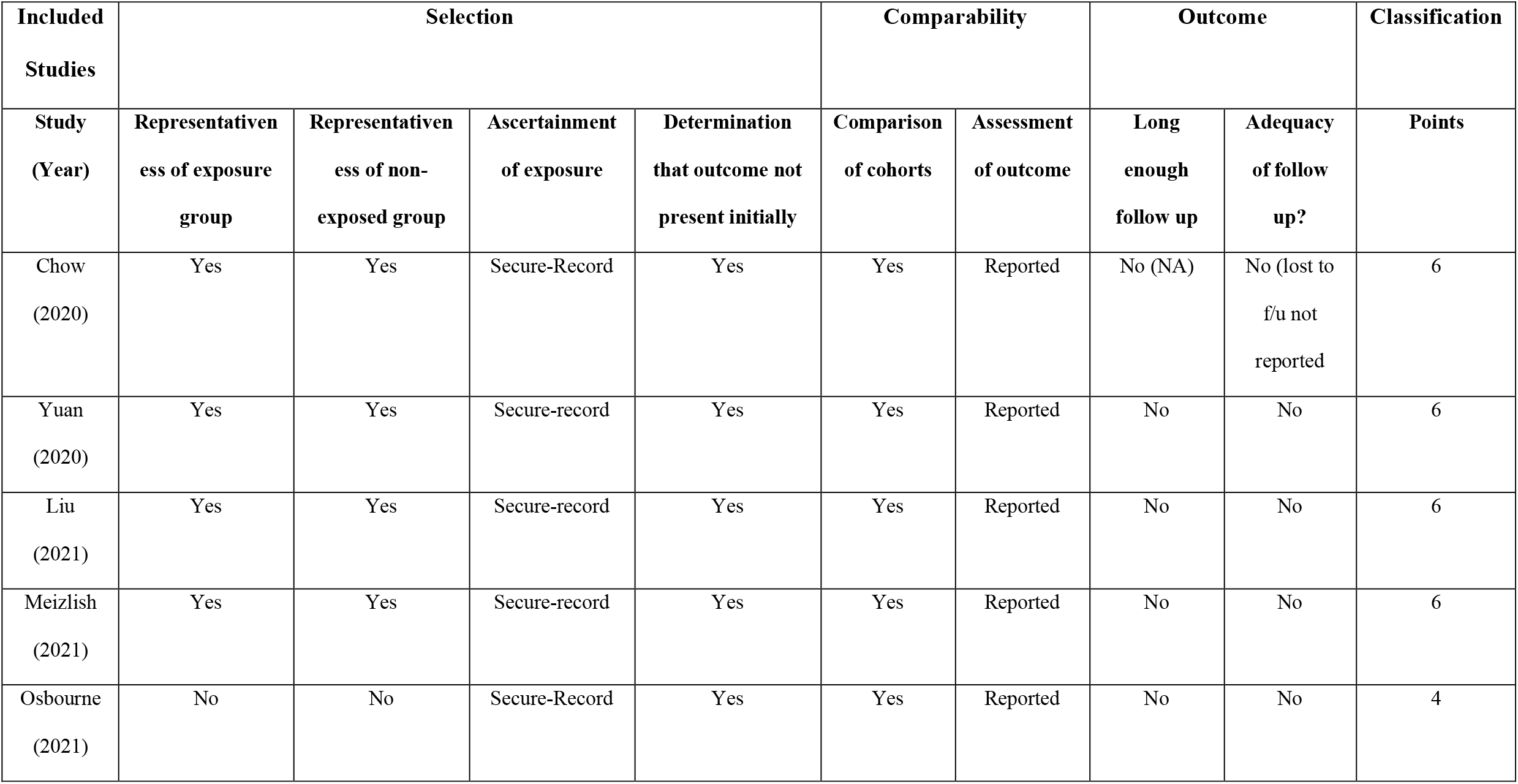
Quality of included studies.

### 3.4 OUTCOME

#### 3.4.1 Overall mortality

There were 6797 participants in the aspirin group and 7268 participants in the no aspirin group. The pooled data from five studies show that the use of aspirin was associated with 53% decrease in overall mortality compared to no aspirin in patients with COVID-19 (adjusted HR 0.47, 95% CI 0.35-0.63, P< 0.001, I^2^= 47%). Figure 2.

**Figure 2:**
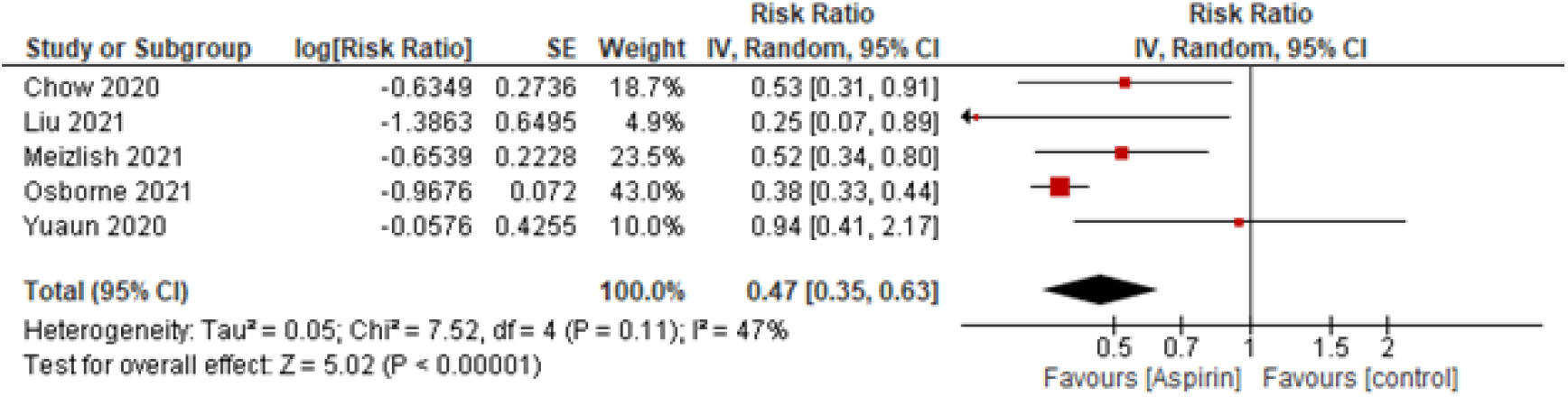
Forest plot of the effect of aspirin use on mortality in adults with COVID-19.

#### 3.4.2 In-hospital mortality

In the analysis restricted to patients hospitalized for COVID-19, the use of aspirin was associated with a 49% reduction in the risk for mortality (HR 0.51, 95% CI 0.33-0.80, P = 0.004, I^2^= 39%). Figure 3.

**Figure 3:**
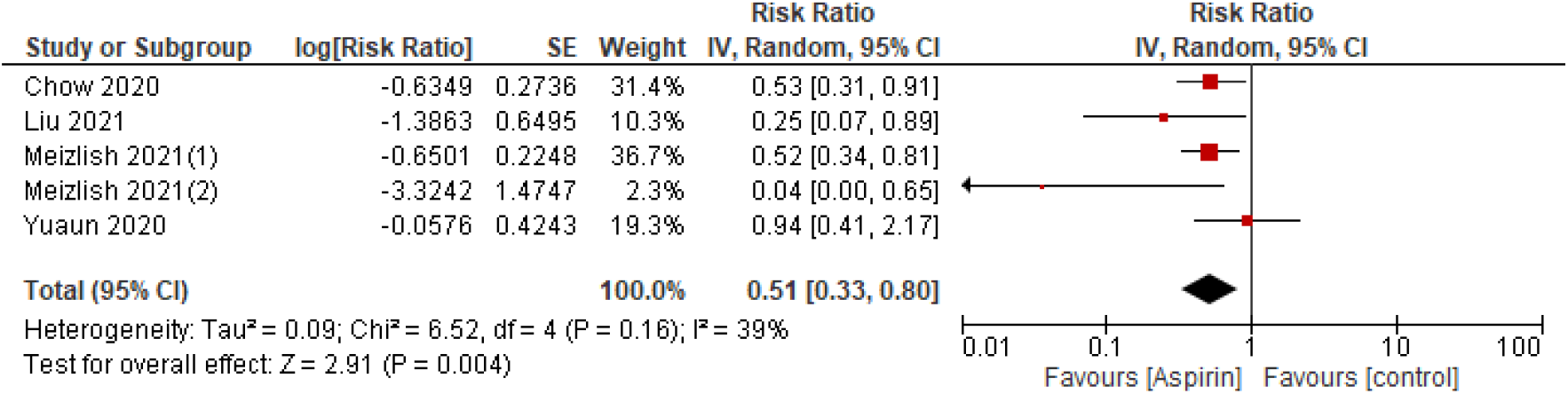
Forest plot of the effect of aspirin use on in-hospital mortality in adults hospitalized with COVID-19.

### 3.5 Sensitivity analysis

The results from sensitivity analysis did not change the overall beneficial effects of aspirin in prevention of mortality in COVID-19 patients.

## 4. Discussion

The pathophysiology of COVID-19 virus in contributing to a prothrombotic and hypercoagulable state is incompletely understood. Studies have implicated the role of severe cytokine storm with marked increase in various cytokines including IL-6, IL-10, TNF-a, and coagulopathy leading to high fatality rates in hospitalized COVID 19 cases^12-14^. While some studies have shown benefits of systemic anticoagulation to reduce mortality in patients who are mechanically ventilated, the benefits of aspirin on mortality are less well known^15^. Aspirin has not only anti-inflammatory and antiplatelet effects but also antiviral effects, which has been shown in vitro and experimental models to reduce replication, propagation, and infectivity of many RNA viruses such as MERS-CoV and CoV-229 E^14-17^. Hence, aspirin was studied as one of the therapeutic options in patients with COVID-19. So far, only a few retrospective studies have investigated the effect of Aspirin on the outcome of mortality in patients with COVID-19. And, there are some interventional studies currently investigating the effect^18^. Salah et al. performed the first meta-analysis investigating the effects of Aspirin on mortality in 1054 COVID-19 patients and showed no association^19^. Our current study is up-to-date study with five retrospective cohort studies. Moreover, we have included both overall mortality and in-hospital mortality as separate outcomes.

Results of our meta-analysis suggest that the use of aspirin may be associated with a mortality benefit in patients with COVID-19. Our results differ from the previous meta-analysis. We included few recently published observational studies. Because available studies which have investigated the effects of aspirin on COVID-19 mortality are observational studies, there is a high likelihood for bias by indication. Furthermore, patients taking aspirin are more likely to have cardiovascular disease, which may place them at higher risk for mortality due to the morbidity alone.

Our study has many other limitations. Firstly, all the studies included in the meta-analysis are observational as no RCTs are available comparing aspirin to no aspirin at this time. There is an increased risk of publication bias and other reporting biases such as selective outcome reporting. Nonetheless, to reduce confounding and selection bias, most of the studies in the analysis used propensity scores. The third limitation is that most studies are single-center studies conducted in China and the United States. One of the studies included patients from VA, who are disproportionately older, and male compared to the general population. It can affect the generalizability of the study to other ethnic groups. Fourth, most of the studies did not mention the dosing of aspirin. The heterogeneity of the studies is also higher with I^2^ of 47% and 39% respectively in overall mortality and in hospital mortality. We did not perform meta regression to adjust for this heterogeneity.

## 5. Conclusion

Our study shows that aspirin is associated with a decrease in both overall mortality and in-hospital mortality in patients with COVID-19. The role of Aspirin in COVID-19 infection should be further examined in a clinical trial. Our evidence is from observational studies with a limited sample size. The limitations of our study include lack of RCTs, inclusion of Cohort studies which have high risk of bias and confounders, and wide confidence interval of individual studies.

## Data Availability

The link of secondary data are on the references of the manuscript.

## 6. AKNOWLEGDEMENT

None

